# Automated CTA-Based Morphometric Analysis for Predicting Primary Entry Tear Location in Stanford Type A Aortic Dissection

**DOI:** 10.64898/2026.07.30.26359380

**Authors:** Xin Fang, Shuang Li, Yongchun You, Wanjiang Li, Xiaobo Zhou, Chunyan Lu, Kang Li, Chaoyi Qin, Kaiyue Diao

**Affiliations:** Department of Radiology, West China Hospital, Sichuan University, Chengdu, China; West China Biomedical Big Data Center, West China Hospital, Sichuan University, Chengdu, China; School of Biomedical Informatics, University of Texas Health Science Center at Houston, Houston, USA; Department of Cardiovascular Surgery and Cardiovascular Surgery Research Laboratory, West China Hospital, Sichuan University, Chengdu, China

**Author notes:** Chaoyi Qin and Kaiyue Diao are corresponding authors. Correspondence: Chaoyi Qin[MD,PHD], Department of Cardiovascular Surgery and Cardiovascular Surgery Research Laboratory, West China Hospital, Sichuan University, No. 37 Guoxue Alley, Wuhou District, Chengdu, Sichuan 610041, China. Kaiyue Diao[MD,PHD], Department of Radiology, West China Hospital, Sichuan University, No. 37 Guoxue Alley, Wuhou District, Chengdu, Sichuan 610041, China. Xin Fang and Shuang Li contributed equally to this study.

**Keywords:** aortic dissection, computed tomography angiography, entry tear, false lumen, machine learning

## Abstract

**Background:** Preoperative localization of the primary entry tear in Stanford type A aortic dissection helps determine the extent of aortic repair but remains challenging on emergency computed tomography angiography (CTA). We developed and externally validated a model to distinguish ascending aortic from arch entry tears using automatically quantified aortic morphology.

**Methods:** This retrospective study included a single-center development cohort of 680 patients, comprising 529 with ascending and 151 with arch entry tears, and an independent ImageTAAD external cohort of 89 patients, comprising 47 with ascending and 42 with arch entry tears. Two three-dimensional nnU-Net models segmented the true lumen, patent false lumen, false lumen thrombus, and anatomical zones on preoperative arterial-phase CTA. We extracted 131 morphological features and used leakage-controlled five-fold cross-validation to select features and compare five classifiers. In the development cohort, intraoperative tear location served as the reference standard, supplemented by CTA review. The final model and deployment threshold were locked before external validation.

**Results:** Five features representing ascending false lumen and thrombus burden, arch geometry, and ascending aortic caliber were retained. Random forest achieved the highest internal cross-validated discrimination, with an area under the receiver operating characteristic curve (AUC) of 0.862 (95% CI, 0.833 to 0.892). At the pooled out-of-fold threshold of 0.281, sensitivity was 92.7% and specificity was 66.5%. In external validation, the locked model achieved an AUC of 0.785 (95% CI, 0.692 to 0.879). At the development-derived threshold of 0.253, sensitivity was 97.6% and specificity was 17.0%. In an exploratory post hoc analysis, a cohort-specific threshold yielded 78.6% sensitivity and 68.1% specificity.

**Conclusions:** A five-feature model automatically derived from preoperative CTA distinguished ascending from arch primary entry tears and retained discrimination externally. Site-specific threshold assessment and prospective multicenter validation are required before clinical use.

## Introduction

Stanford type A aortic dissection (TAAD) is a cardiovascular emergency associated with high in-hospital mortality, particularly among patients managed without surgery (1, 2). Together with the anatomical extent of dissection, branch involvement, patient risk, and local expertise, primary entry tear location informs repair strategy(3). Tear location is also associated with clinical presentation and outcomes(4). Surgical resection of the primary entry tear is the standard strategy for TAAD. An ascending aortic entry tear may be managed with ascending aortic or hemiarch replacement, whereas an arch entry tear may favor more extensive repair, such as total arch replacement with or without a frozen elephant trunk. Although many factors contribute to the selection of surgical strategy, extended arch repair entails longer operative times and may increase selected perioperative morbidities, while comparative studies have not consistently shown higher early mortality(5–8). Accurate preoperative identification of primary entry tear location is therefore essential for surgical planning and risk stratification.

Preoperative computed tomography angiography (CTA) is the principal imaging modality for evaluating TAAD and identifying the primary entry tear. However, accurate localization of the entry tear remains a challenge in clinical practice (accuracy ranging from 64% to 88.0% across physicians with varying levels of seniority and clinical experience, as reported by Kim et al.(9)). In addition, in emergency imaging, where non-gated CTA is more commonly used, diagnostic confidence and inter-reader agreement can be further reduced by pulsation artifacts at the ascending aortic dissection(10, 11). A surgical perspective summarized accurate preoperative CT entry-tear identification rates of 40%–100% across earlier series(12). Automated methods hole promise but offer limited remedy. AI applications in aortic dissection have largely targeted lumen and anatomical segmentation, while entry-tear segmentation in TAAD remains substantially more challenging(13).

An alternative approach is to infer the location of the primary entry tear indirectly from aortic morphology rather than by directly visualizing the entry tear. Mechanistically, the primary entry tear directly governs false lumen hemodynamics and determines which aortic zones and branch vessels are preferentially affected, thereby encoding its spatial information within the overall dissection morphology. Takami et al.(14) provided early support for this principle, showing that CT morphological parameters including pericardial effusion, aortic diameter, and false lumen thrombosis were associated with primary entry tear location; however, they reported only odds ratios without classification accuracy, leaving the clinical utility of such an approach unestablished. Although entry-tear segmentation remains challenging, automated segmentation of the true lumen, false lumen, and anatomical regions provides a basis for quantitative morphological analysis that still requires feature-level validation(13, 15). These features, obtainable without directly visualizing the entry tear, may serve as objective surrogates for entry-tear localization.

Therefore, we develop and validate a machine learning model that predicts whether the primary entry tear in TAAD is located in the ascending aorta or the aortic arch in origin from quantitative morphological features automatically extracted from preoperative CTA segmentations, using intraoperative findings as the reference standard. Rather than detecting the entry tear directly, we sought to infer its location from the morphological imprint it leaves on the dissected aorta.

## Methods

### Study Design and Patient Selection

This study used two independent cohorts. The development cohort came from a single tertiary center and was used for model training and internal five-fold cross-validation. The external validation cohort came from the publicly available ImageTAAD database. The development cohort was assembled retrospectively and approved by the institutional review board of West China Hospital (No. 2025 Review [1706]). Informed consent was waived given the retrospective design. We screened consecutive patients admitted with acute aortic dissection for eligibility. Patients were eligible if they had a preoperative arterial-phase CTA and a primary entry tear in the ascending aorta or aortic arch (Stanford type A). We excluded patients whose entry tear was confined to the descending aorta, whose primary entry tear location could not be determined, or whose image quality was inadequate for analysis. The reference standard was the entry tear location confirmed intraoperatively by the attending cardiac surgeon, supplemented by preoperative CTA review.

A total of 873 eligible patients were identified at our center. To prevent data leakage, the earliest 100 consecutive patients were used to train the segmentation model and were excluded from downstream analysis. After further exclusion for inadequate segmentation quality, the final development cohort comprised 680 patients, 529 with an ascending entry tear and 151 with an arch entry tear. Applying the same eligibility criteria to the ImageTAAD database yielded 89 patients for external validation, 47 with an ascending and 42 with an arch entry tear (arch prevalence 47.2%). The patient selection process for both cohorts is shown in Figure 1.

**Figure 1.**
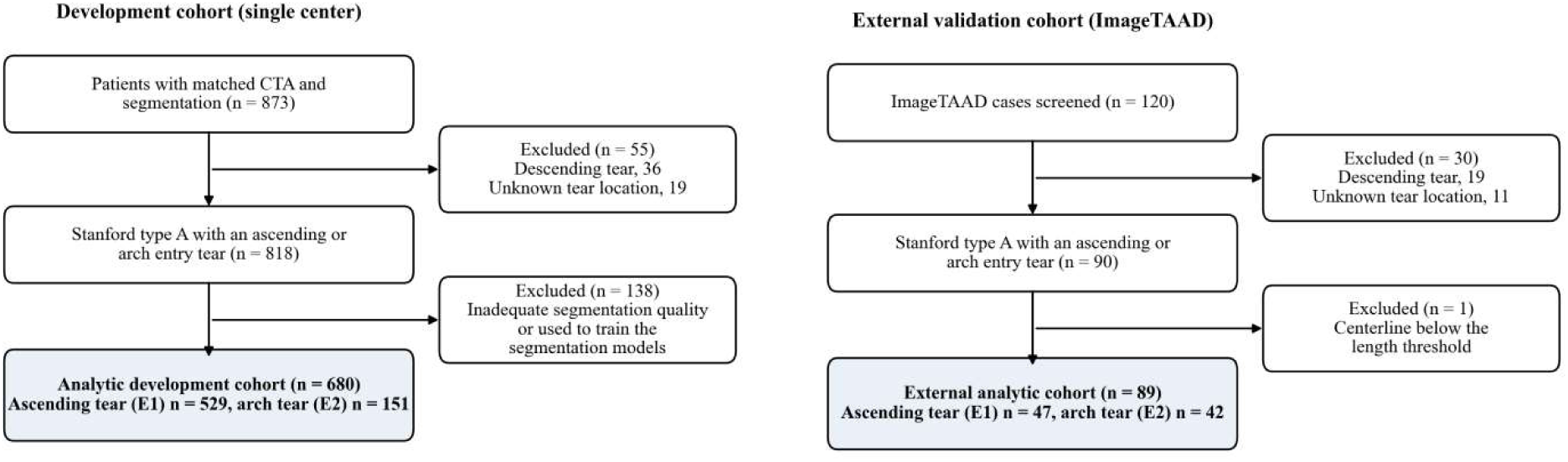
Patient selection flow diagram. Flow of the development cohort (single center) and the external validation cohort (ImageTAAD) through screening, exclusion, and analysis. The development cohort yielded 680 analytic patients (529 ascending and 151 arch entry tears) and the external cohort yielded 89 analytic patients (47 ascending and 42 arch entry tears).

### CTA Image Acquisition

Preoperative CTA was performed on three scanner platforms: United Imaging uCT 960+ and uCT 780 (371 patients), Siemens SOMATOM Definition Flash and Force (208 patients), and GE Revolution CT and Apex (101 patients). Tube voltage was 120 kVp in 462 patients and 100 kVp in 208 patients, with a small number acquired between 70 and 140 kVp. Arterial-phase images were reconstructed with vendor soft-tissue kernels at a median slice thickness of 1.0 mm (range 0.5 to 2.0) and a median in-plane pixel spacing of 0.81 mm (Supplementary Table S1). All examinations used iodinated contrast with a standard arterial-phase bolus-tracking protocol.

### Automated Aortic Segmentation

Two nnU-Net models, configured in 3D full-resolution mode, were used to segment the arterial-phase CTA. The first model produced four labels: background (0), true lumen (TL, 1), patent false lumen (FL, 2), and false lumen thrombus (FLT, 3). The second model produced 24 labels following the SVS/STS zone classification, comprising 10 aortic trunk zones (Zone 0 through Zone 9) and 13 branch vessels (the innominate, left common carotid, and left subclavian arteries; the celiac and superior mesenteric arteries; and the bilateral renal, common iliac, internal iliac, and external iliac arteries).

Both models were trained on the 100 reserved earliest cases, which were manually annotated by two cardiovascular radiologists with 7 years of experience, using the default five-fold cross-validation of the nnU-Net pipeline. The lumen model reached a mean Dice coefficient of 0.93 for the true lumen, 0.89 for the false lumen, and 0.38 for thrombus. The lower thrombus value reflected its small and variable volume. The zone model reached a mean Dice of 0.89 (Supplementary Methods 1). The trained models were then applied to the 680 analytic patients for fully automated inference

### Centerline Extraction and Cross-sectional Sampling

Aortic centerlines were extracted from the zone segmentation masks using VMTK 1.5.0 (VTK 9.2.6). Each mask was first converted to a surface mesh by marching cubes (scikit-image 0.24.0, iso-level 0.5). The mesh was then smoothed by Taubin filtering (passband 0.1, 30 iterations) and, for meshes exceeding 20,000 triangles, decimated to 10% of the original face count. Centerlines were finally computed with the Voronoi-based algorithm and automated seed selection. The resulting centerlines were resampled at 1.0-mm intervals. In all 680 patients, they met the predefined quality thresholds for the aortic trunk, namely a minimum length of 150 mm with at least 50 points for the trunk and 5 points for each branch vessel. Detailed mesh-processing parameters are provided in Supplementary Methods 2.

Cross-sectional profiles were sampled along the trunk centerline at 2-mm intervals on planes perpendicular to the local tangent. Sampling used a radius of three times the local trunk radius and nearest-neighbor interpolation. Because the automated centerline consistently began distal to the aortic valve annulus, we substituted axial slices at the proximal Zone 0 root in all 680 patients. We also substituted axial slices at the distal Zone 9 extreme in the 376 patients whose centerline ended proximal to the distal segmentation boundary. The aortic arch, where ascending and arch entry tears are distinguished, was always covered by perpendicular sampling.To align the anatomical boundary with the surgical reference standard, we redefined the ascending-to-arch boundary at the innominate artery origin rather than at the conventional SVS/STS Zone 0 to Zone 1 junction. The innominate origin was located by identifying innominate voxels adjacent to Zone 0 by 26-connectivity and projecting their centroid onto the trunk centerline.

At this landmark, Zone 0 was divided into a proximal ascending segment (TEM class E1) and a distal portion. The distal portion was merged with Zones 1 and 2 to form the arch segment (TEM class E2). This correction succeeded in 673 of 680 patients (99.0%). In the remaining 7 patients without innominate labeling, the original SVS/STS boundary was retained. Morphological features were then computed at two anatomical granularities: fine-zone features following the original SVS/STS definitions, and coarse-region features based on the TEM-corrected boundaries. This allowed feature selection to retain the more informative boundary definition.

### Quantitative Feature Extraction

A total of 131 quantitative features were extracted per patient and organized into 11 groups (Supplementary Table S2). The complete false lumen was defined as the union of patent FL (label 2) and thrombosed false lumen (FLT, label 3). Six groups (82 features) captured zone-level morphology, and five groups (49 features) characterized centerline-level spatial profiles.

The six zone-level groups were as follows. Group 1 (24 features) described regional false lumen volume distribution across both fine zones and TEM-corrected regions. Group 2 (16 features) described false lumen area gradients within the ascending and arch segments. Group 3 (10 features) measured true lumen compression as the FL/(FL + TL) ratio at each cross-sectional level. Group 4 (10 features) described thrombus distribution, including regional thrombosis ratios and the FLT center of mass. Group 5 (12 features) described global aortic morphology, such as total volume, segment lengths, curvature, tortuosity, mean diameter, and maximum inscribed sphere radius. Group 6 (10 features) described false lumen involvement of seven major branch vessels above a 5% luminal threshold.

The five centerline-level groups were as follows. Group 7 (9 features) described the false lumen area profile along the full centerline, including its gradient, the position of first appearance and of peak area, and the spatial skewness and kurtosis of the distribution. Group 8 (16 features) described per-region gradient statistics across the four TEM-corrected segments. Group 9 (4 features) described the phase-transition points at which the false lumen ratio first exceeded 30% and 50% of the total lumen area. Group 10 (5 features) described true lumen compression dynamics. Group 11 (15 features) described false lumen curve shape, captured by five Fourier coefficient amplitudes and ten normalized histogram bin means. All positional features were normalized to the range 0 to 1, and full feature definitions are provided in Supplementary Table S2.

Six features had missing values in more than 5% of patients, arising from structural anatomical conditions rather than from extraction errors. For example, the FLT center of mass along the centerline is mathematically undefined in patients without a thrombosed false lumen and was recorded as missing. All remaining features were extracted successfully in all 680 patients.

### Statistical Analysis

Feature selection reduced the initial 131-feature set through a five-stage pipeline. It first removed near-zero-variance features, then pruned the lower-AUC feature from each correlated pair (|r| > 0.85), then applied Boruta analysis, then stability selection by LASSO, and finally retained the features confirmed by both methods. Five classifiers were then trained under five-fold stratified cross-validation: elastic net, LASSO, random forest, XGBoost, and LightGBM. Class imbalance was addressed by class weighting, missing values were imputed within each fold using training-set medians, and the threshold was set by the Youden index. With 151 arch entry tear events and five final predictors, the events-per-variable ratio was approximately 30, which exceeds the conventional minimum for stable model fitting. The classifier with the highest cross-validated AUC was selected as the final model. Feature-selection details are provided in Supplementary Tables S3A and S3B, and model hyperparameters in Supplementary Table S4.

Discrimination was assessed by AUC with pairwise DeLong tests, calibration by calibration plots, the Brier score, calibration slope and intercept, and the Spiegelhalter Z test, and clinical utility by decision curve analysis. Model interpretability was examined with SHAP values. The final model, with all preprocessing and the operating threshold locked from the internal data, was then externally validated on the ImageTAAD cohort with the pipeline applied unchanged, and between-cohort distribution shift was quantified by the Kolmogorov-Smirnov statistic and the population stability index. Continuous variables were compared by the Mann-Whitney U test or the Student t test, and categorical variables by the chi-square or Fisher exact test, at a two-sided alpha of 0.05. Analyses were performed in Python 3.13 and R 4.4.3, and reporting was assessed against the TRIPOD+AI guideline (Supplementary Checklist).

## Results

### Study Population

The analytic cohort comprised 680 patients, 529 (77.8%) with an ascending entry tear (E1) and 151 (22.2%) with an arch entry tear (E2) (Table 1). Patients with an arch entry tear were more often male (86.8% versus 71.6%) and current or former smokers (52.3% versus 35.7%, both p < 0.001), and had a marginally higher body mass index (25.6 versus 25.0 kg/m2, p = 0.046). The two groups were comparable in age, hypertension, diabetes, Marfan syndrome, acute presentation, and bovine aortic arch.

**Table 1.** Baseline characteristics stratified by primary entry-tear location.

| Variable | Ascending (n=529) | Arch (n=151) | p-value |
| --- | --- | --- | --- |
| Age (years), median [IQR] | 51.00 [44.00, 59.00] | 51.00 [41.50, 58.00] | 0.257 |
| BMI (kg/m<sup>2</sup>), mean (SD) | 24.95 (3.63) | 25.63 (3.82) | 0.046 |
| Male sex, n (%) | 379 (71.6) | 131 (86.8) | <0.001 |
| Hypertension, n (%) | 367 (69.4) | 111 (73.5) | 0.364 |
| Diabetes, n (%) | 13 (2.5) | 4 (2.6) | 1.000 |
| Marfan syndrome, n (%) | 18 (3.4) | 1 (0.7) | 0.092 |
| Smoking, n (%) | 189 (35.7) | 79 (52.3) | <0.001 |
| Acute presentation, n (%) | 460 (87.0) | 135 (89.4) | 0.487 |
| Bovine aortic arch, n (%) | 11 (2.1) | 0 (0.0) | 0.135 |

Data are median [IQR], mean (SD), or n (%). P values: Mann–Whitney U test or Student t test, as appropriate, for continuous variables; chi-square test or Fisher exact test, as appropriate, for categorical variables.
IQR, interquartile range; SD, standard deviation; BMI, body mass index.

### Automated Segmentation and Morphological Differences

Automated segmentation, centerline extraction, and feature computation completed successfully in all 680 patients. Representative automated segmentation overlays are shown in Figure 2. In the validated folds, the lumen model reached a mean Dice coefficient of 0.93 for the true lumen, 0.89 for the false lumen, and 0.38 for thrombus, and the zone model reached a mean Dice of 0.89 (Supplementary Methods 1).

**Figure 2.**
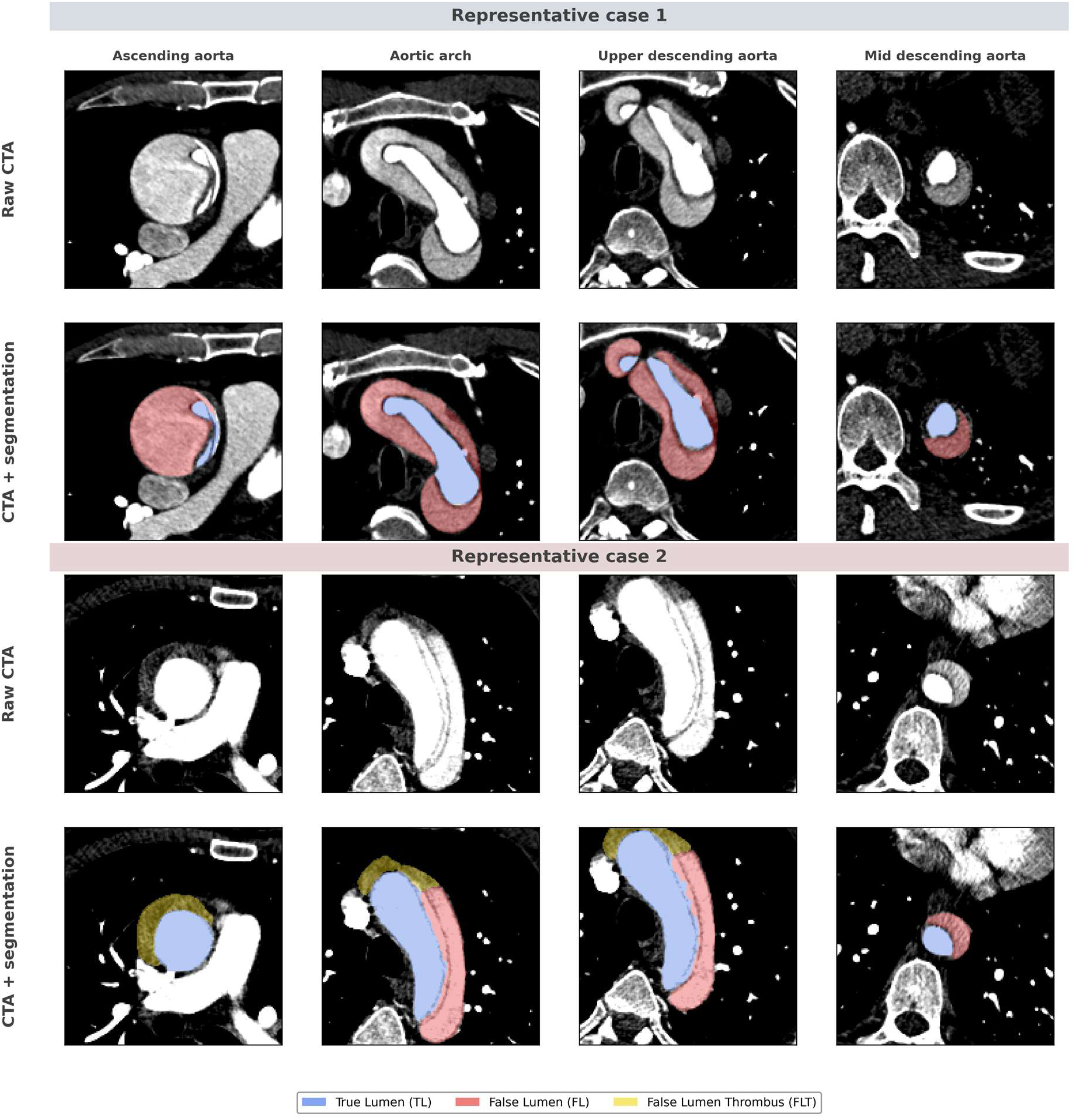
Automated aortic segmentation across anatomical zones in representative cases. Axial CTA images from two representative cases (case 1, upper block; case 2, lower block) are shown as paired raw images and automated nnU-Net segmentation overlays at the ascending aorta, aortic arch, upper descending aorta, and mid descending aorta. Periwinkle indicates the true lumen, rose the false lumen, and gold the thrombosed false lumen. The segmentation model was applied without manual correction.

Representative false lumen area profiles along the aortic centerline are shown in Figure 3 and demonstrate substantial within-group heterogeneity.

**Figure 3.**
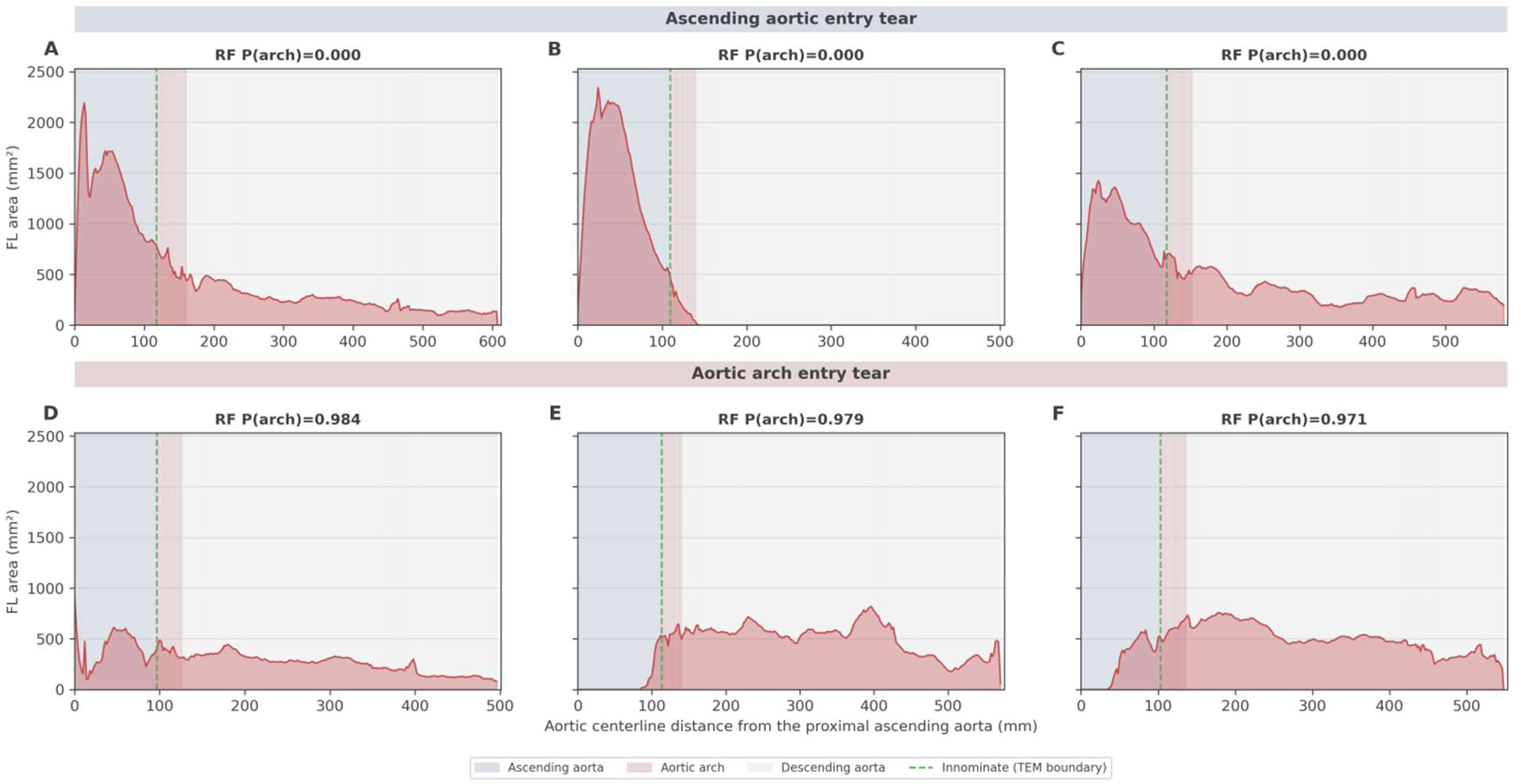
False lumen centerline profile patterns in ascending versus arch entry tear. Panels A–C show ascending aortic entry-tear cases, and panels D–F show aortic arch entry-tear cases. Longitudinal false lumen cross-sectional area profiles plotted against aortic centerline distance from the proximal ascending aorta for three representative patients with ascending aortic entry tears (E1, upper row) and three with aortic arch entry tears (E2, lower row). The green dashed line marks the innominate artery origin, which is the TEM boundary. Background shading denotes the aortic segments, with blue-gray for the ascending aorta, rose for the aortic arch, and light gray for the descending aorta. The value above each panel is the out-of-fold random forest probability of an arch entry tear, shown as RF P(arch).

These patterns were mirrored in the selected features (Figure 4B). Compared with patients with an ascending entry tear, those with an arch entry tear had a lower ascending false lumen ratio (median 0.29 versus 0.43), a larger arch maximum inscribed radius (15.7 versus 14.8 mm) and arch mean diameter (38.4 versus 36.3 mm), a smaller ascending maximum inscribed radius (17.8 versus 19.1 mm), and a higher ascending thrombosis ratio (0.34 versus 0.00). All differences were significant (p < 0.001). A comparison of selected morphological features between the two groups is provided in Supplementary Table S5.

**Figure 4.**
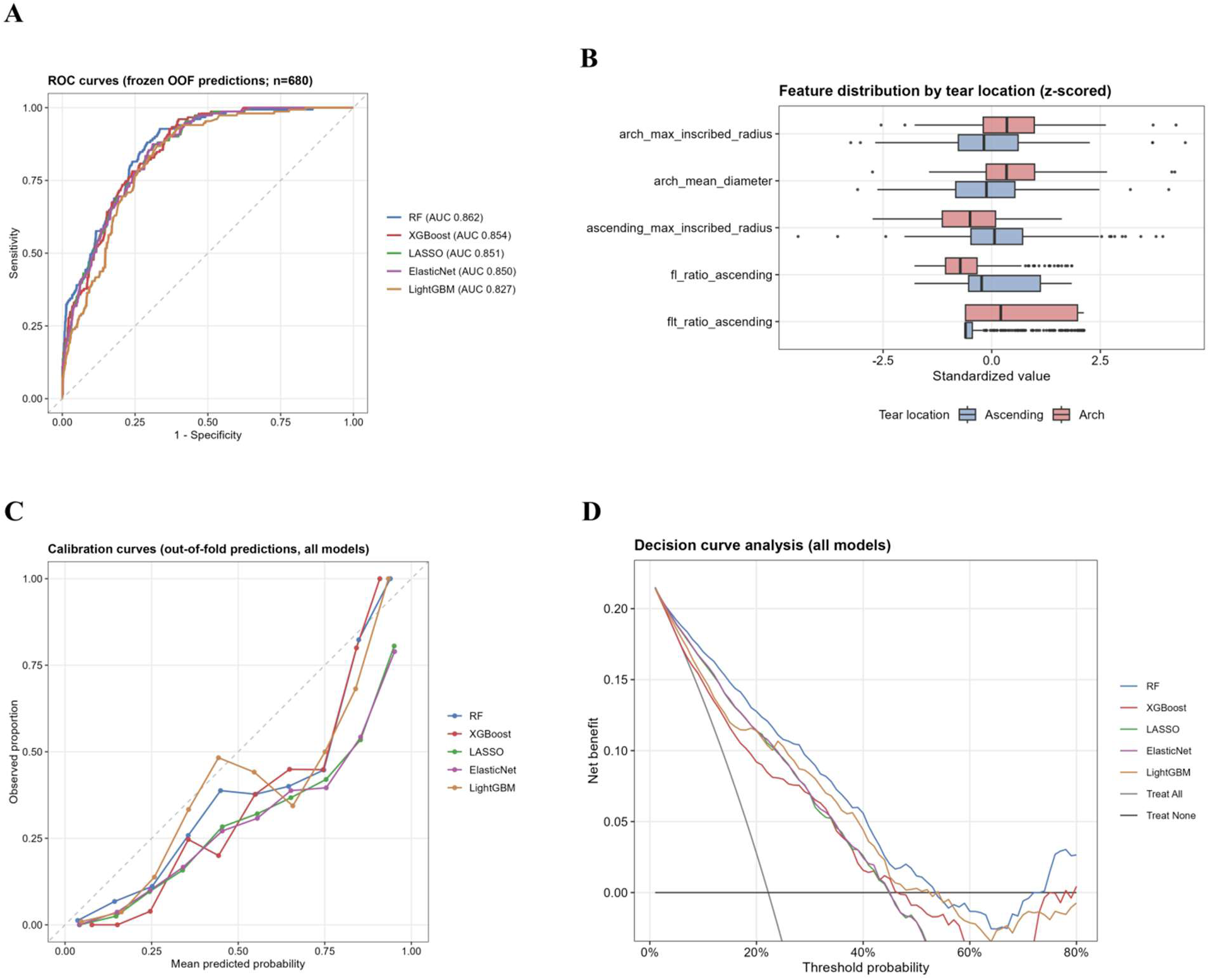
Model discrimination, feature distribution, calibration, and clinical utility. (A) Receiver operating characteristic curves for all five classifiers under five-fold stratified cross-validation (n = 680, 5 features), with area under the curve values shown in parentheses. (B) Standardized feature distributions by entry tear location for the five selected features. (C) Calibration curves based on out-of-fold predictions for all five classifiers, where the dashed diagonal represents perfect calibration. (D) Decision curve analysis showing the net benefit of all five classifiers relative to the treat-all and treat-none strategies across threshold probabilities of 0 to 80%.

### Feature Selection and Model Performance

The five-stage pipeline reduced the initial 131 features to 5, each with a selection stability of at least 99% across bootstrap iterations (Supplementary Table S6). The retained features captured three aspects of dissection morphology: ascending false lumen and thrombus burden (fl_ratio_ascending and flt_ratio_ascending), arch geometry (arch_max_inscribed_radius and arch_mean_diameter), and ascending aortic caliber (ascending_max_inscribed_radius). All five classifiers were trained on this common feature set. The random forest achieved the highest discrimination, with an AUC of 0.862 (95% CI 0.833 to 0.892) and, at the Youden-optimal threshold, a sensitivity of 92.7% and a specificity of 66.5% (Table 2). It significantly outperformed LightGBM (p = 0.004), whereas its advantage over XGBoost, LASSO, and elastic net was not significant (p = 0.174, 0.256, and 0.230, respectively). The random forest was therefore selected as the final model.

**Table 2.** Discrimination of candidate models in internal cross-validation.

| Model | AUC (95% CI) | Sensitivity, % | Specificity, % | Threshold | P vs RF |
| --- | --- | --- | --- | --- | --- |
| RF | 0.862 (0.833–0.892) | 92.7 | 66.5 | 0.281 | Reference |
| XGBoost | 0.854 (0.824–0.883) | 96.0 | 60.1 | 0.331 | 0.174 |
| LASSO | 0.851 (0.821–0.881) | 87.4 | 68.4 | 0.418 | 0.256 |
| Elastic Net | 0.850 (0.820–0.880) | 87.4 | 69.0 | 0.420 | 0.230 |
| LightGBM | 0.827 (0.795–0.860) | 86.8 | 67.5 | 0.297 | 0.004 |

Internal performance from 5-fold cross-validation (pooled out-of-fold predictions). AUC 95% CI by the DeLong method.
P vs RF: DeLong test for the difference in AUC versus the random forest (reference model).
Operating threshold by the Youden index. AUC, area under the ROC curve; RF, random forest.

### Model Calibration and Clinical Utility

The final model was well calibrated (Figure 4). The Brier score was 0.131 and the calibration slope was 1.032, close to the ideal of 1.0. The calibration intercept was −0.724, indicating slight systematic overestimation of predicted probabilities. The Spiegelhalter Z test showed no significant miscalibration (Z = −0.914, p = 0.361). Decision curve analysis showed positive net benefit relative to both the treat-all and treat-none strategies across a clinically relevant range of threshold probabilities (Figure 4).

### Model Interpretability

SHAP analysis of the final random forest model ranked fl_ratio_ascending as the dominant predictor (mean |SHAP| = 0.108), followed by arch_max_inscribed_radius (0.100), flt_ratio_ascending (0.089), ascending_max_inscribed_radius (0.072), and arch_mean_diameter (0.069) (Figure 5, Supplementary Table S6). These predictors drew on ascending lumen and thrombus burden (Groups 1 and 4) and arch geometry (Group 5). A higher ascending false lumen ratio and a larger ascending caliber shifted the prediction toward an ascending entry tear (E1). A larger arch inscribed radius and mean diameter, together with a higher proportion of thrombus in the ascending false lumen, shifted it toward an arch entry tear (E2).

**Figure 5.**
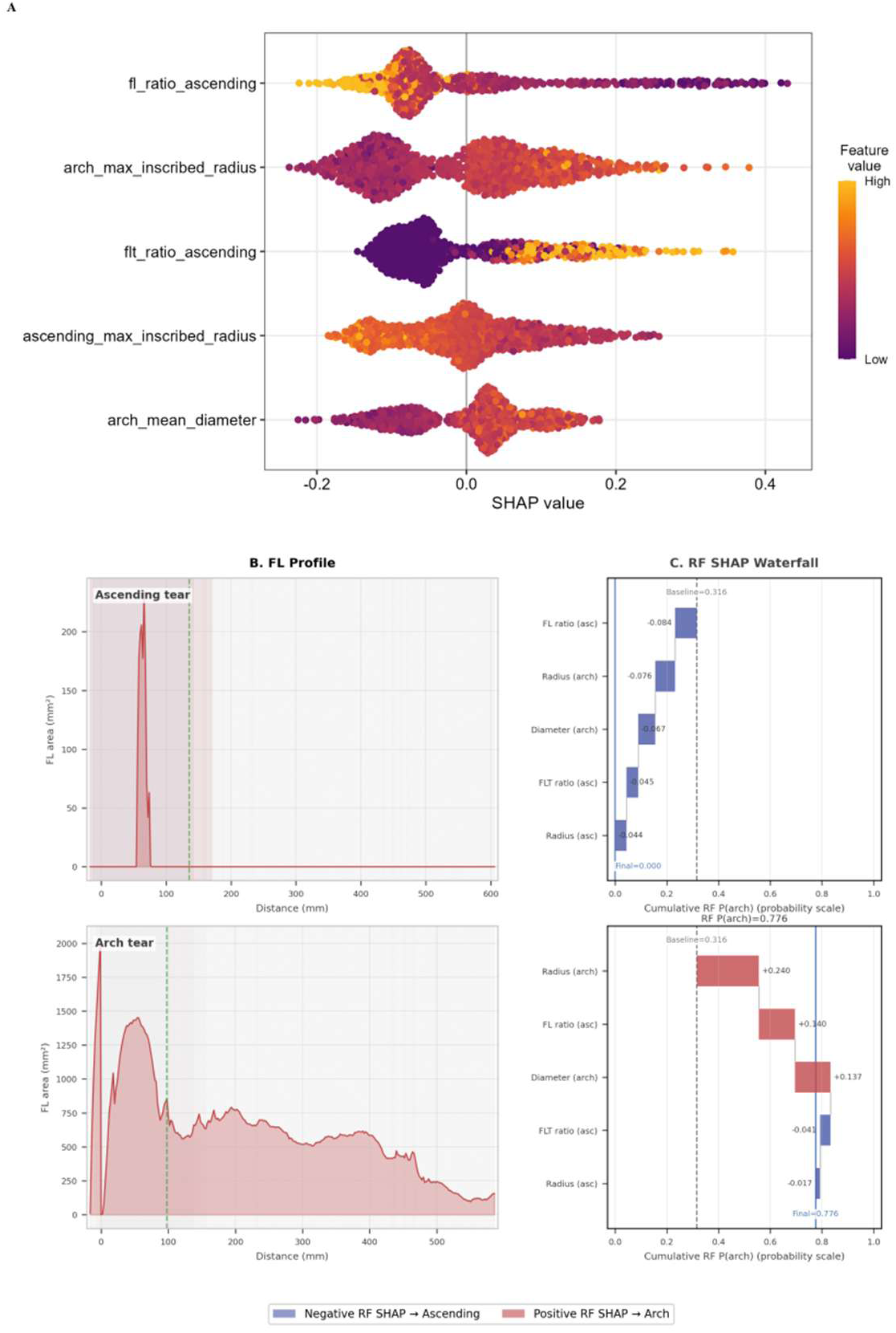
SHAP-based model interpretability and representative case illustrations. (A) SHAP beeswarm plot derived from the final random forest model trained on the full dataset. Each point represents one patient. Horizontal position indicates the SHAP value, that is the direction and magnitude of the contribution toward an arch entry tear prediction, and color indicates the standardized feature value, with yellow for high and purple for low. (B–C) Representative case illustrations for a typical ascending entry tear patient (upper, RF P(arch) = 0.000) and a typical arch entry tear patient (lower, RF P(arch) = 0.776). (B) False lumen cross-sectional area along the aortic centerline with anatomical-region background shading; the green dashed line marks the innominate artery origin. (C) Probability-scale RF SHAP waterfall plots showing how the five selected features shift the baseline probability to the patient-specific RF P(arch).

### External Validation

The external cohort comprised 89 patients, of whom 42 had an arch entry tear (prevalence 47.2%). The locked random forest retained moderate discrimination, with an AUC of 0.785 (95% CI 0.692 to 0.879) (Table 3). At the internally locked threshold, the model was highly sensitive but poorly specific (sensitivity 97.6%, specificity 17.0%, accuracy 55.1%). A threshold re-optimized on the external cohort recovered balanced operating characteristics (sensitivity 78.6%, specificity 68.1%, accuracy 73.0%). This indicates that the loss at the locked threshold reflected a shift in the predicted-probability distribution rather than a loss of separability.

**Table 3.** Internal and external performance of the final random forest model, with an exploratory post hoc threshold analysis.

| **Metric** | **Internal cross-validation** | **External (locked threshold)** | **External exploratory analysis (post hoc cohort-specific threshold)** |
| --- | --- | --- | --- |
| No. of patients | 680 | 89 | 89 |
| No. with arch tear | 151 | 42 | 42 |
| Operating threshold | 0.281 | 0.253 | 0.501 |
| AUC (95% CI) | 0.862 (0.833–0.892) | 0.785 (0.692–0.879) | 0.785 (0.692–0.879) |
| Sensitivity, % | 92.7 | 97.6 | 78.6 |
| Specificity, % | 66.5 | 17.0 | 68.1 |
| PPV, % | 44.2 | 51.2 | 68.8 |
| NPV, % | 97.0 | 88.9 | 78.0 |
| Accuracy, % | 72.4 | 55.1 | 73.0 |
| F1 score | 0.598 | 0.672 | 0.733 |
| Brier score | 0.131 | 0.188 | 0.188 |
| Calibration slope | 1.032 | 1.392 | 1.392 |
| Calibration intercept | -0.724 | -0.240 | -0.240 |

Data are from the final random forest model. Internal performance was estimated using pooled out-of-fold predictions from five-fold cross-validation. External validation was performed in the independent ImageTAAD cohort of 89 patients.
The internal operating threshold of 0.281 was selected using the Youden index based on pooled out-of-fold predictions. The locked deployment threshold of 0.253 was selected using out-of-bag predictions from the final random forest model refit on the complete development cohort and was applied unchanged to the external cohort. Both thresholds were derived exclusively from development data. Their difference reflects the use of different internal estimation procedures and does not represent threshold adjustment using external data.
The post hoc cohort-specific threshold of 0.501 was selected using the Youden index and evaluated in the same external cohort. The corresponding sensitivity, specificity, PPV, NPV, accuracy, and F1 score are therefore optimistic and are reported only as an exploratory analysis of the effect of threshold selection. They should not be interpreted as independently validated performance.
AUC, Brier score, calibration slope, and calibration intercept are threshold-independent and are therefore identical across the two external operating points. The 95% CIs for AUC were calculated using the DeLong method.
AUC indicates area under the receiver operating characteristic curve; CI, confidence interval; NPV, negative predictive value; PPV, positive predictive value.

Calibration degraded accordingly, with a Brier score of 0.188 and a calibration slope of 1.392. The slope above 1.0 showed that predicted probabilities were less dispersed than observed risk. All five selected features differed significantly between cohorts (Kolmogorov-Smirnov statistic 0.19 to 0.68, all p ≤ 0.007), and population stability indices indicated major distribution shift (0.36 to 5.37). The shift was driven chiefly by a systematically larger external aorta. The arch mean diameter was 45.9 versus 36.9 mm, the arch inscribed radius 18.6 versus 15.1 mm, and the ascending inscribed radius 22.1 versus 19.0 mm (Supplementary Table S7). Discrimination nonetheless remained well above chance.

## Discussion

In this study we developed and externally validated a model that predicts primary entry tear location in TAAD from the morphological footprint that the entry tear leaves on the dissected aorta. Using five interpretable features extracted automatically after segmentation of routine preoperative CTA, the model separated ascending from arch entry tears with an area under the curve of 0.862 (95% CI 0.833 to 0.892) on internal cross-validation and 0.785 (95% CI 0.692 to 0.879) on an independent external cohort. The automatic pipeline may offer a localization approach that fits the emergency workflow.

Efforts to localize the primary entry tear preoperatively have so far taken three forms, and each has met an obstacle. The first is direct visual interpretation of CT, which is strongly experience-dependent. In Kim et al.’s(9) retrospective multi-reviewer study of 50 patients with acute TAAD, surgeons showed lower and less consistent accuracy than cardiovascular radiologists in predicting intimal tear location (75.0±7.7% vs. 86.7±1.2%). Notably, sensitivity for arch tears was particularly low, averaging only 62.4±12.5% across all reviewers and 59.7±11.5% among surgeons. The second is an indirect, morphology-based route that infers the entry tear site from the shape of the dissection rather than from the entry tear itself. Takami et al. (14)linked regional caliber and false-lumen thrombosis to entry location, whereas Furui et al.(16) related tear characteristics to false-lumen patency; both were single-center retrospective association studies without classifier performance or external validation. The third is the most direct automated route, deep-learning segmentation of the entry tear, which has fared worst of all. In the ImageTAAD benchmark, entry-tear segmentation was reported as substantially more challenging than segmentation of the major aortic structures(13).

Our approach combined the strengths of these two morphological and deep-learning routes while avoiding the obstacle that had limited each. Deep-learning studies have achieved high accuracy for true- and false-lumen segmentation across type A and type B dissection datasets, whereas entry-tear and thrombus segmentation and clinical feature extraction remain more variable and task-dependent(13, 17–19). We therefore used it for what it does well and built the prediction on that substrate rather than on the entry tear. On this foundation we quantified the false lumen along the entire centerline and reduced the dissection to five anatomically defined measurements. The SHAP analysis showed that the prediction rested on quantities a clinician already reasons with. A false lumen concentrated in a dilated ascending aorta pointed to an ascending entry tear, and a wider arch pointed to an arch entry tear, with each entry tear enlarging the segment that receives its outflow. The thrombus feature was less intuitive, because a greater share of ascending false lumen thrombosis favored an arch entry tear. Flow may contribute to this finding because an arch entry tear could permit retrograde filling of the ascending false lumen, where slow recirculating flow may promote thrombosis. This hypothesis is only indirectly consistent with a three-patient computational study of type B dissection, including two patients after TEVAR, in which shear rate and residence time were incorporated into predictions of false-lumen thrombosis(20).

The five selected features also matched the variable families that Takami had identified manually, namely regional caliber and false lumen thrombosis(14). This convergence may lend mechanistic support to our data-driven result. The contrast with other machine-learning work in dissection is also informative. Prior machine-learning studies have predicted postoperative adverse outcomes or mortality mainly from clinical variables, whereas CT radiomics has achieved high discrimination for dissection detection using high-dimensional features that are less directly anatomical(21–23). Our model instead draws its accuracy from a handful of interpretable anatomical measurements. This may be the main contribution of the present work, an entry-tear localization tool that is automated, mechanistically transparent, and quantitatively validated.

Our external validation results demonstrated that the model’s discriminative ability was preserved in an independent external cohort acquired from a different clinical setting, with an AUC of 0.785, although its internally fixed operating point did not transfer directly. The probability scale shifted, so the internally fixed threshold labeled almost every external patient as an arch entry tear. When the threshold was re-set on the external cohort, sensitivity and specificity returned to a balanced level. Preserved discrimination despite poor threshold transportability suggests a calibration problem; however, the appropriate update, whether threshold or intercept adjustment, logistic recalibration, or model revision, should be evaluated in an adequately sized target cohort(24). The larger aortic dimensions in the external cohort may have contributed to the observed probability shift, but this hypothesis requires further study. External implementation should therefore include site-specific assessment of calibration and operating thresholds before clinical use. Nonetheless, prospective, multicenter validation remains necessary to confirm its value in real-world clinical practice.

It is still a hot topic about arch strategies for TAAD. Hemiarch replacement was considered as the standard approach for patients with ATAAD while extensive arch repair becomes more common due to its better long-term outcomes(25). One essential criteria for arch strategy selection is aortic arch pathology. A primary entry tear in the arch is the key indication for extensive arch repair in TAAD(26). Wherefore, feasible and automated prediction of the primary entry tear in this emergency workflow can facilitate the selection of surgical strategy and accelerate the preparation of the whole team, including anesthetist, perfusionist and surgical nurses.

This study has several limitations. First, we addressed the binary choice between an ascending and an arch entry tear in surgically treated type A dissection. Other presentations, including primary entry tears of descending origin, were outside the present scope and remain for future work. Second, the model was developed at a single center and tested on one external dataset of modest size, and the external confidence interval was correspondingly wide, so multicenter validation is needed before routine use. Third, we evaluated discrimination, calibration, and net benefit, but we did not assess whether use of the model changes surgical decisions or patient outcomes, which will require prospective study.

In summary, a small set of morphological features, derived automatically from routine preoperative CTA, can localize the primary entry tear in type A aortic dissection to the ascending aorta or the arch. By inferring the primary entry tear from its morphological footprint, the approach provides a reproducible CTA-derived prediction that could support time-sensitive preoperative surgical planning. Clinical adoption requires prospective multicenter validation and site-specific assessment of the operating threshold.

## Data Availability

The deidentified data supporting the findings of this study are not publicly available because the clinical and imaging data are subject to patient privacy, institutional ethics, and data-governance restrictions. Deidentified data may be made available from the corresponding authors upon reasonable request, subject to institutional approval, review of the proposed research purpose, and execution of an appropriate data-use agreement.The publicly available ImageTAAD dataset used for external validation can be accessed through the link provided below.

https://www.kaggle.com/datasets/xiaoweixumedicalai/imagetaad

## Disclosures

None.

## Author Contributions

Xin Fang: Conceptualization, Methodology, Investigation, Data curation, Software development, Formal analysis, Visualization, Writing - review & editing.

Shuang Li: Conceptualization, Methodology, Investigation, Data curation, Formal analysis, Funding acquisition, Writing - review & editing.

Yongchun You: Data acquisition, Investigation, Clinical evaluation, Writing - review & editing.

Wanjiang Li: Data acquisition, Investigation, Clinical evaluation, Supervision, Writing - review & editing.

Xiaobo Zhou: Methodology, Software development, Algorithmic guidance, Validation, Writing - review & editing.

Chunyan Lu: Clinical evaluation, Investigation, Validation, Writing - review & editing.

Kang Li: Software development, Algorithmic support, Methodology, Validation, Writing - review & editing.

Chaoyi Qin: Conceptualization, Clinical data collection, Clinical expertise, Validation, Funding acquisition, Writing - review & editing.

Kaiyue Diao: Conceptualization, Supervision, Project administration, Methodology, Validation, Writing - review & editing.

## Funding Information

This research was supported by the National Natural Science Foundation of China (No. 82470386) and the Sichuan Provincial Department of Science and Technology (Grant No. 2025ZNSFSC1921).

## Data Sharing Statement

Deidentified data generated or analyzed during this study are not publicly available because of patient privacy and institutional restrictions. Reasonable requests for access to deidentified aggregate data may be directed to the corresponding author, subject to institutional approval.

## Abbreviations

AUC: area under the receiver operating characteristic curve
CTA: computed tomography angiography
FL: false lumen
FLT: false lumen thrombus
KS: Kolmogorov-Smirnov
LASSO: least absolute shrinkage and selection operator
LightGBM: Light Gradient Boosting Machine
NPV: negative predictive value
OOF: out-of-fold
PPV: positive predictive value
PSI: population stability index
RF: random forest
SHAP: Shapley additive explanations
SVS/STS: Society for Vascular Surgery/Society of Thoracic Surgeons
TAAD: type A aortic dissection
TEM: type, entry site, and malperfusion
TL: true lumen
VMTK: Vascular Modeling Toolkit
XGBoost: extreme gradient boosting

## Notes

### Competing Interest Statement

The authors have declared no competing interest.

### Author Declarations

The development cohort was assembled retrospectively and approved by the institutional review board of West China Hospital (No. 2025 Review [1706]). Informed consent was waived given the retrospective design.

## References

1. Hagan PG, Nienaber CA, Isselbacher EM, Bruckman D, Karavite DJ, Russman PL, et al. The International Registry of Acute Aortic Dissection (IRAD): new insights into an old disease. JAMA. 2000;283(7):897–903.

2. Pape LA, Awais M, Woznicki EM, Suzuki T, Trimarchi S, Evangelista A, et al. Presentation, Diagnosis, and Outcomes of Acute Aortic Dissection: 17-Year Trends From the International Registry of Acute Aortic Dissection. J Am Coll Cardiol. 2015;66(4):350–8.

3. Isselbacher EM, Preventza O, Hamilton Black J, 3rd, Augoustides JG, Beck AW, Bolen MA, et al. 2022 ACC/AHA Guideline for the Diagnosis and Management of Aortic Disease: A Report of the American Heart Association/American College of Cardiology Joint Committee on Clinical Practice Guidelines. Circulation. 2022;146(24):e334–e482.

4. Takahashi B, Kamohara K, Morokuma H, Yunoki J, Kawaguchi A. Impact of primary entry tear locations on outcomes in acute type A aortic dissection. Sci Rep. 2025;15(1):13981.

5. Ma WG, Zhang W, Wang LF, Zheng J, Ziganshin BA, Charilaou P, et al. Type A aortic dissection with arch entry tear: Surgical experience in 104 patients over a 12-year period. J Thorac Cardiovasc Surg. 2016;151(6):1581–92.

6. Salem M, Friedrich C, Rusch R, Frank D, Hoffmann G, Lutter G, et al. Is total arch replacement associated with an increased risk after acute type A dissection? J Thorac Dis. 2020;12(10):5517–31.

7. Poon SS, Theologou T, Harrington D, Kuduvalli M, Oo A, Field M. Hemiarch versus total aortic arch replacement in acute type A dissection: a systematic review and meta-analysis. Ann Cardiothorac Surg. 2016;5(3):156–73.

8. Nappi F, Gambardella I, Singh SSA, Salsano A, Santini F, Spadaccio C, et al. Survival following acute type A aortic dissection: a multicenter study. J Thorac Dis. 2023;15(12):6604–22.

9. Kim JS, Park KH, Lim C, Kim DJ, Jung Y, Shin YC, et al. Prediction of Intimal Tear Site by Computed Tomography in Acute Aortic Dissection Type A. Korean Circ J. 2016;46(1):48–55.

10. Budeanu RG, Broemmer C, Budeanu AR, Pop M. Comparing the Diagnostic Performance of ECG Gated versus Non-Gated CT Angiography in Ascending Aortic Dissection: A GRRAS Study. Tomography. 2022;8(5):2426–34.

11. Nagpal P, Agrawal MD, Saboo SS, Hedgire S, Priya S, Steigner ML. Imaging of the aortic root on high-pitch non-gated and ECG-gated CT: awareness is the key! Insights Imaging. 2020;11(1):51.

12. Appoo JJ, Pozeg Z. Strategies in the surgical treatment of type A aortic arch dissection. Ann Cardiothorac Surg. 2013;2(2):205–11.

13. Song S, Qiu H, Huang M, Zhuang J, Lu Q, Shi Y, et al. Domain knowledge based comprehensive segmentation of Type-A aortic dissection with clinically-oriented evaluation. Med Image Anal. 2025;102:103512.

14. Takami Y, Tajima K, Kato W, Fujii K, Hibino M, Munakata H, et al. Can we predict the site of entry tear by computed tomography in patients with acute type a aortic dissection? Clin Cardiol. 2012;35(8):500–4.

15. Isensee F, Jaeger PF, Kohl SAA, Petersen J, Maier-Hein KH. nnU-Net: a self-configuring method for deep learning-based biomedical image segmentation. Nat Methods. 2021;18(2):203–11.

16. Furui M, Uesugi N, Matsumura H, Hayashida Y, Kuwahara G, Fujii M, et al. Relationship between false lumen morphology and entry tear in acute type A aortic dissection. Eur J Cardiothorac Surg. 2024;65(2).

17. Cao L, Shi R, Ge Y, Xing L, Zuo P, Jia Y, et al. Fully automatic segmentation of type B aortic dissection from CTA images enabled by deep learning. Eur J Radiol. 2019;121:108713.

18. Xiang D, Qi J, Wen Y, Zhao H, Zhang X, Qin J, et al. ADSeg: A flap-attention-based deep learning approach for aortic dissection segmentation. Patterns (N Y). 2023;4(5):100727.

19. Zhuang C, Wu Y, Qi Q, Zhao S, Sun Y, Hou J, et al. A Fully Automatic Pipeline of Identification, Segmentation, and Subtyping of Aortic Dissection from CT Angiography. Cardiovasc Eng Technol. 2025;16(4):465–80.

20. Menichini C, Cheng Z, Gibbs RGJ, Xu XY. A computational model for false lumen thrombosis in type B aortic dissection following thoracic endovascular repair. J Biomech. 2018;66:36–43.

21. Luo H, Liu X, Yang Y, Tang B, He P, Ding L, et al. Preoperative prediction of major adverse outcomes after total arch replacement in acute type A aortic dissection based on machine learning ensemble. Sci Rep. 2025;15(1):20930.

22. Zhang J, Xiong W, Yang J, Sang Y, Zhen H, Tan C, et al. Enhanced machine learning models for predicting one-year mortality in individuals suffering from type A aortic dissection. J Thorac Cardiovasc Surg. 2025;169(4):1191–200 e3.

23. Guo Y, Chen X, Lin X, Chen L, Shu J, Pang P, et al. Non-contrast CT-based radiomic signature for screening thoracic aortic dissections: a multicenter study. Eur Radiol. 2021;31(9):7067–76.

24. Van Calster B, McLernon DJ, van Smeden M, Wynants L, Steyerberg EW, Topic Group ‘Evaluating diagnostic t, et al. Calibration: the Achilles heel of predictive analytics. BMC Med. 2019;17(1):230.

25. From the American Association of Neurological Surgeons ASoNC, Interventional Radiology Society of Europe CIRACoNSESoMINTESoNESOSfCA, Interventions SoIRSoNS, World Stroke O, Sacks D, Baxter B, et al. Multisociety Consensus Quality Improvement Revised Consensus Statement for Endovascular Therapy of Acute Ischemic Stroke. Int J Stroke. 2018;13(6):612–32.

26. Ma WG, Chen Y, Chen SW, Zhang W, Zheng J, Li QG, et al. Frozen elephant trunk for acute type A aortic dissection: long-term outcomes over two decades. Eur Heart J. 2026;47(20):2452–65.

